# Detection of *Salmonella* Typhi bacteriophages in surface waters as a scalable approach to environmental surveillance

**DOI:** 10.1101/2023.02.14.23285806

**Authors:** Sneha Shrestha, Kesia Esther Da Silva, Jivan Shakya, Alexander T. Yu, Nishan Katuwal, Rajeev Shrestha, Mudita Shakya, Sabin Bikram Shahi, Shiva Ram Naga, Christopher LeBoa, Kristen Aiemjoy, Isaac I. Bogoch, Senjuti Saha, Dipesh Tamrakar, Jason R. Andrews

**Author notes:** **Correspondence:** Kesia Esther da Silva, Division of Infectious Diseases and Geographic Medicine, Stanford University School of Medicine, Biomedical Innovations Building, Room 3600, Stanford, CA 94025, USA. Contributed equally (Sneha Shrestha, Kesia Esther Da Silva). Contributed equally (Dipesh Tamrakar, Jason R. Andrews).

## Abstract

Environmental surveillance, using detection of *Salmonella* Typhi DNA, has emerged as a potentially useful tool to identify typhoid-endemic settings; however, it is relatively costly and requires molecular diagnostic capacity. We sought to determine whether *S*. Typhi bacteriophages are abundant in water sources in a typhoid-endemic setting, using low-cost assays. We collected drinking and surface water samples from urban, peri-urban and rural areas in 4 regions of Nepal. We performed a double agar overlay with *S*. Typhi to assess the presence of bacteriophages. We isolated and tested phages against multiple strains to assess their host range. We performed whole genome sequencing of isolated phages, and generated phylogenies using conserved genes. *S*. Typhi-specific bacteriophages were detected in 54.9% (198/361) of river water samples and 6.3% (1/16) drinking water samples from the Kathmandu Valley and Kavrepalanchok. Water samples collected within or downstream of population-dense areas were more likely to be positive (72.6%, 193/266) than those collected upstream from population centers (5.3%, 5/95) (p=0.005). In urban Biratnagar and rural Dolakha, where typhoid incidence is low, only 6.7% (1/15, Biratnagar) and 0% (0/16, Dolakha) samples contained phages. All *S*. Typhi phages were unable to infect other *Salmonella* and non-*Salmonella* strains, nor a Vi-knockout *S*. Typhi strain. Representative strains from *S*. Typhi lineages were variably susceptible to the isolated phages. Phylogenetic analysis showed that *S*. Typhi phages belonged to two different viral families (*Autographiviridae* and *Siphoviridae*) and clustered in three distinct groups. *S*. Typhi bacteriophages were highly abundant in surface waters of typhoid-endemic communities but rarely detected in low typhoid burden communities. Bacteriophages recovered were specific for *S*. Typhi and required Vi polysaccharide for infection. Screening small volumes of water with simple, low-cost plaque assays enables detection of *S*. Typhi phages and should be further evaluated as a scalable tool for typhoid environmental surveillance.

**Highlights:**

- Typhoid phages are detectable in surface water using simple assays, in communities with high typhoid burden.
- Bacteriophages are highly specific for *S*. Typhi and required Vi polysaccharide for infection.
- *S*. Typhi phages have a broad lytic activity against the *S*. Typhi strains circulating in Nepal.
- Phage plaque assay can be used as a low-cost tool to identify communities where typhoid is endemic.
- The high abundance of phages in river water suggest that this could be an alternative to molecular methods for environmental surveillance for typhoid.

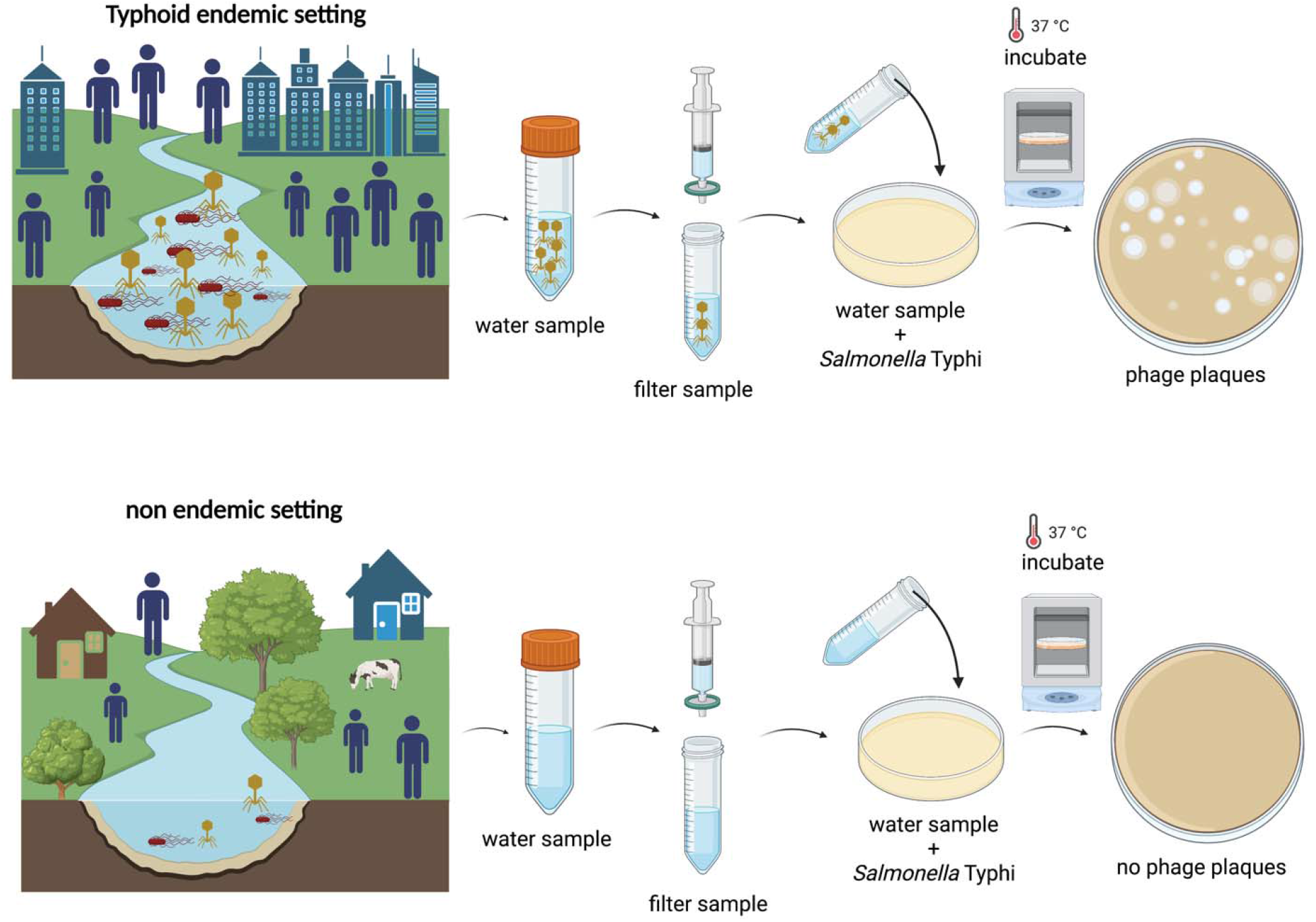

## 1. Introduction

Typhoid fever is a bacterial infection caused by the Gram-negative bacterium *Salmonella enterica* subspecies enterica serovar Typhi (*S*. Typhi) and is usually contracted by ingestion of contaminated food or water (1). An estimated 11 million cases of typhoid fever occur annually worldwide causing more than 100,000 deaths each year (2). The highest burden occurs in low- and middle-income countries (LMICs) with poor access to improved water supply and sanitation (3). Successful management of typhoid fever using antibiotics is becoming increasingly difficult due to drug resistance (4). The continued emergence and rapid expansion of antimicrobial resistance to *S*. Typhi have spurred accelerated efforts to roll out new conjugate vaccines for the prevention of typhoid infection in high-risk populations (5).

Routine typhoid fever vaccination use has been very limited in the past, even in endemic areas. Recently the World Health Organization (WHO) recommended the introduction of typhoid conjugate vaccines (TCV) in countries with the highest burden of typhoid disease or a high burden of antimicrobial-resistant *S*. Typhi (6). However, the scarcity of data on typhoid burden in many at-risk countries poses a major hurdle to the adoption of TCV in national immunization programs (7). Currently, the estimates of typhoid fever burden have been generated via short-term population-based research studies, and these data are frequently geographically and temporally limited (8). Given the current lack of data and the challenges faced in population-based surveillance, environmental surveillance is emerging as an important tool in the fight against typhoid fever (9).

Previous studies have shown environmental surveillance as a tool to identify high-risk settings for enteric fever transmission (8–10). Despite the clear role of contaminated water in the transmission of typhoid, detection of the organisms has proven challenging, with *S*. Typhi difficult to isolate by culture methods from water and other environmental samples (8). Molecular detection of *S*. Typhi DNA is now the primary method for detecting *S*. Typhi in the environment, but requires molecular laboratory infrastructure and trained staff, and estimated costs are hundreds of dollars per sample (11).

Bacteriophages (hereinafter referred to as phages) are viruses that infect bacteria. Their abundance and host specificity make them a promising alternative or complimentary tool for the environmental monitoring of pathogenic bacteria (12). *S*. Typhi phages have been characterized for decades, comprised the major typing system for the bacteria (based on their susceptibility to phage panels), and were used clinically as treatment prior to the discovery of chloramphenicol. One study from 1930 found that *S*. Typhi phages were abundant and seasonal in surface waters in Kolkata (13). However, few studies have characterized the distribution and abundance of *S*. Typhi phages in typhoid-endemic settings or examined their genetic diversity.

In this study, we investigated the presence and abundance of typhoid phages in surface and drinking water across diverse communities in Nepal with differing population density and typhoid burden. We hypothesized that *S*. Typhi phages would be readily detectable, using simple, low-cost assays, in communities with high typhoid burden and not frequently detected in communities with lower typhoid burden.

## 2. Methods

### 2.1. Bacterial strains, Vi phages and culture conditions

A complete list of strains used in this study can be found in Table S1. The attenuated *Salmonella enterica* serovar Typhi strain BRD948 was used for propagation. This strain is not only heavily capsulated but is also attenuated and can be used in a containment level 2 environment (14). Bacteria were cultured using Tryptic Soy Broth (TSB) (HiMedia Laboratories, USA), at 37 °C with aeration, supplemented with agar as required. The optical density of liquid culture was adjusted to a 0.5 McFarland standard. Phages used in this paper as a control for the experiments were isolated from clinical samples and correspond to a *S*. Typhi Vi phage collection, designated types II (*Siphoviridae)* and types III, IV, V, VI, VII (*Podoviridae*). Vi phages were obtained from Cambridge University, Cambridge, United Kingdom, but the original sources of these phages date from the 1930s through 1955 (15).

### 2.2. Sampling sites

Between November 2019 and May 2022, environmental water samples were collected from the Kathmandu Valley and three other districts of Nepal (Kavrepalanchok, Dolakha and Morang). We collected river water samples from 5 major rivers and a combined river segment in Kathmandu Valley and Kavrepalanchok on a monthly basis from November 2019 to July 2021 (except in April and May 2020 due to government-imposed lockdown during the COVID-19 pandemic). A total of 19 different sites, 16 in Kathmandu valley and 3 in Kavrepalanchok, each separated by approximately 5 km, located upstream and downstream from the river confluence were sampled. This sampling strategy let us sample the upstream sites with relatively sparse settlement, the densely populated central settlement area and also downstream sites mostly located around agricultural areas. Within Kathmandu valley, the majority of the sites (10/16) were either located in rivers inside, around or downstream to Kathmandu Metropolitan city, the capital city of Nepal, with a population density of ∼17,103 people per km^2^. Unmanaged sewage, waste disposal and lack of their proper regulation in this highly dense area has led to their direct disposal into the river system in Kathmandu. We also sampled a river flowing through Banepa, a peri-urban city in Kavrepalanchok district just outside the Kathmandu Valley, which similarly has untreated waste and sewage disposal into the river system. Drinking water samples were collected from both Kathmandu and Kavre, from tap (direct supply from municipal storage or stored in tanks, bought from vendors outside Kathmandu) or ground-well of 16 different households. Previous studies have demonstrated a very high incidence of typhoid in both communities (7).

In Dolakha, sampling was performed in the month of October, 2021, in sparsely populated, mostly rural village areas where the population density was significantly less and there was minimal or no visible sewage contamination. All samples collected were from river water around settlement areas, with only a few sites selected being farther from human settlement. A previous study in Dolakha found typhoid to be an uncommon cause of acute febrile illness (16). Biratnagar, in Morang district in Southeastern Nepal, is another metropolitan city in Nepal with a population density of ∼3200 per km^2^. Samples were collected from river sites upstream, around, and downstream of the settlement area, from man-made river canals which were originally created for irrigation purposes and from tap or tube wells of households that used it for drinking along with other purposes. Most of the tube well samples were collected from poor slum communities. No studies of typhoid incidence or prevalence among febrile patients have been reported from Biratnagar. In our review of blood culture results from three largest hospitals (one government and two private hospitals) in the city, *S*. Typhi was very rarely recovered, and clinicians reported that typhoid cases were relatively uncommon.

### 2.3. Sample collection and filtrate preparation

We collected 50 mL samples of water from the surface, against river current, in a sterile bottle and placed them into separate bags on ice and transported them to the laboratory for further processing. We performed a turbidity test using a densitometer (DEN-1, Grant Instruments Ltd., England), and samples were passed through a 0.22 µm syringe filter (Axiva, India) to obtain the filtrate that was used for evaluating the presence of phages against *S*. Typhi. The filtered sample was stored at 4^°^C until further use.

At each sampling site, team members collected information on abiotic factors that may affect bacterial or viral survival, including presence of sewage pipes, open drain water carrying liquid and solid waste, and potential fecal exposure. Water pH, temperature, and oxygen reduction potential were also evaluated using Apera Instruments AI311. Team members also recorded whether humans or animals were interacting with the water and the type of interaction that was observed. All information was recorded in REDCap (v5.19.15).

### 2.4. Bacteriophage isolation and amplification

Phage screening was performed by a double agar overlay method (17). For each sample, 1 ml of filtrate was added to 100 µl of overnight liquid bacterial culture and incubated at room temperature for 10 min to allow phage absorption. Molten Tryptic Soy Agar (0.7% agar) (3 mL) was added to the filtrate-bacteria mix and poured over solid hard nutrient agar base. Plates were incubated at 37□ overnight and the presence of phages resulted in the production of visible plaques (zones of lysis) in a confluent lawn of the host bacterium. Individual plaques were selected from screening plates based on their morphology and size. Plaques were picked using sterile tips and resuspended in 200 µl SM buffer. Soft agar overlay lawn culture of *S*. Typhi was prepared and 10 µl of plaque-SM buffer solution was streaked across the *S*. Typhi lawn culture to obtain isolated plaques. Only isolated plaques were processed further. To obtain high concentration stocks of isolated phages, we performed propagation assays using the double agar overlay method described above. Following overnight incubation clear zones were scraped and immersed in a 5ml SM buffer and kept for one hour at 4^°^C. The solution was then centrifuged at 4,000 rpm for 20 min and the supernatant was passed through a 0.22 µm syringe filter to obtain a sterile phage stock solution. Finally, the titer of stock solution was obtained by plating serial dilutions of the stock in a confluent lawn of the host bacterium and counting visible plaques to determine the approximate ratio of plaque-forming units per milliliter (PFU mL^-1^).

### 2.5. Determination of phage host range

The lytic activity and specificity of phages was determined by screening each against different host bacterium strains using the standard double agar overlay method previously described. Serial dilutions of each phage stock were prepared and spotted on *S*. Typhi to obtain a concentration where plaques could be observed. Dilutions where plaques could be distinguished were used to spot phages on *S*. Typhi CT18, *S*. Typhi Ty2, *S*. Typhi Ty2 ΔVi (Vi capsule knockout mutant Δ*tviB*), *S*. Typhi Δ*fliC* (flagellate knockout mutant), *Salmonella* enterica serovar Paratyphi A (ATCC: 9150), *Salmonella* Typhimurium LT2, *Escherichia coli* (ATCC: 25922), *Pseudomonas aeruginosa* (ATCC: 27853), *Staphylococcus aureus* (ATCC: 25923) and *Klebsiella pneumoniae* (clinical isolate). To test diversity and activity spectra of phages across different *S*. Typhi lineages, phage stocks were also tested against 37 *S*. Typhi clinical isolates obtained from a surveillance study comprising different genotypes circulating in Nepal.

### 2.6. Whole-genome sequencing

We randomly selected 27 phages from our collection to perform whole genome sequencing. Bacteriophage DNA was extracted using the Phage DNA Isolation Kit (Norgen Biotek Corp., Thorold, ON, Canada) according to the manufacturer’s instructions. The purity and concentration of the DNA were determined using a Qubit 4 fluorometer (Invitrogen, Qubit). Whole genome sequencing was performed using the Illumina Miseq platform (Illumina, San Diego, CA, USA) to generate paired-end reads of 300 bp in length. Sequence data quality was checked using FastQC v0.11.9 to remove low quality reads (18). We summarized all quality indicators using MultiQC v1.7 (19). Species identification was confirmed with Kraken2 (20). Genome assembly was performed using SPAdes Genome Assembler v.3.15.2 and assemblies that resolved as one contig were included in the analysis. Finally, the genomes were annotated using the RAST server (Rapid Annotation using Subsystem Technology) (https://rast.nmpdr.org/rast.cgi) with subsequent manual curation.

### 2.7. Phylogenetic analysis

The annotation of complete phage genomes was also manually inspected using Geneious version 11.0.12 (Biomatters Ltd) to screen for the presence of phage-related regions containing structural proteins. The complete sequences were analyzed adjusting the direction of nucleotide sequences. Phylogenetic trees were generated using selected concatenated phage signature genes. All genomes were screened for the presence of genes encoding terminase and tail fiber structures. We constructed alignments using the program ClustalW version 2.1 with default parameters. Phylogenetic tree was inferred using the Maximum Likelihood method with RAxML v8.2.10. Bootstrapping was set to 100 replicates and the tree rooted. The resulting phylogenies were visualized and annotated using the iTOL v5 online version (21). An additional 27 fiber tail protein sequences and 27 terminase protein sequences from GenBank were included in the phylogenetic analysis for comparison. We selected *Caudovirales* representative phage genomes that were matched with our collection using IDseq (https://czid.org/). The genomes that matched our sequences with avarage percent-identity higher than 90% were included in the analysis. The additional 27 bacteriophage genomes selected for study were isolated from five different host genus: *Citrobacter sp*., *Escherichia sp*., *Klebsiella sp*., *Salmonella spp*., and *Shigella sp*., from different geographical locations.

### 2.8. Data availability

Whole genome sequencing data was submitted to GenBank under BioProject accession number PRJNA933946.

## 3. Results

### 3.1. Sampling and phage detection

A total of 428 environmental water samples were collected from four different regions in Nepal (Table 1; Figure 1). Between November 2019 and October 2021, we collected 377 samples from Kathmandu and Kavrepalanchok. These samples included river water (n = 361) from 19 sites and drinking water (n = 16) from 16 households (Figure 1b). *S*. Typhi phages were present in 54.8% (198/361) of the total river samples and 6.3% (1/16) of the drinking water samples. Most samples (72.6%, 193/266) collected within or downstream of the densely populated parts of the city contained *S*. Typhi phages, while only 5.3% (5/95) of samples from the sites upstream to the population contained these phages. In addition, the upstream sites had lower concentrations of phages than the downstream. No seasonal pattern was observed for phage detection (Figure S1). In September 2021, a total of 16 river water samples were collected from Dolakha (Figure 1c), and none contained *S*. Typhi phages. In May 2022, a total of 35 environmental water samples were also collected from Biratnagar. These samples included river (n = 7), canal (n = 8), and drinking water (n = 20). No *S*. Typhi phages were detected in river or drinking water; one sample from canal water contained *S*. Typhi phages.

**Table 1.**
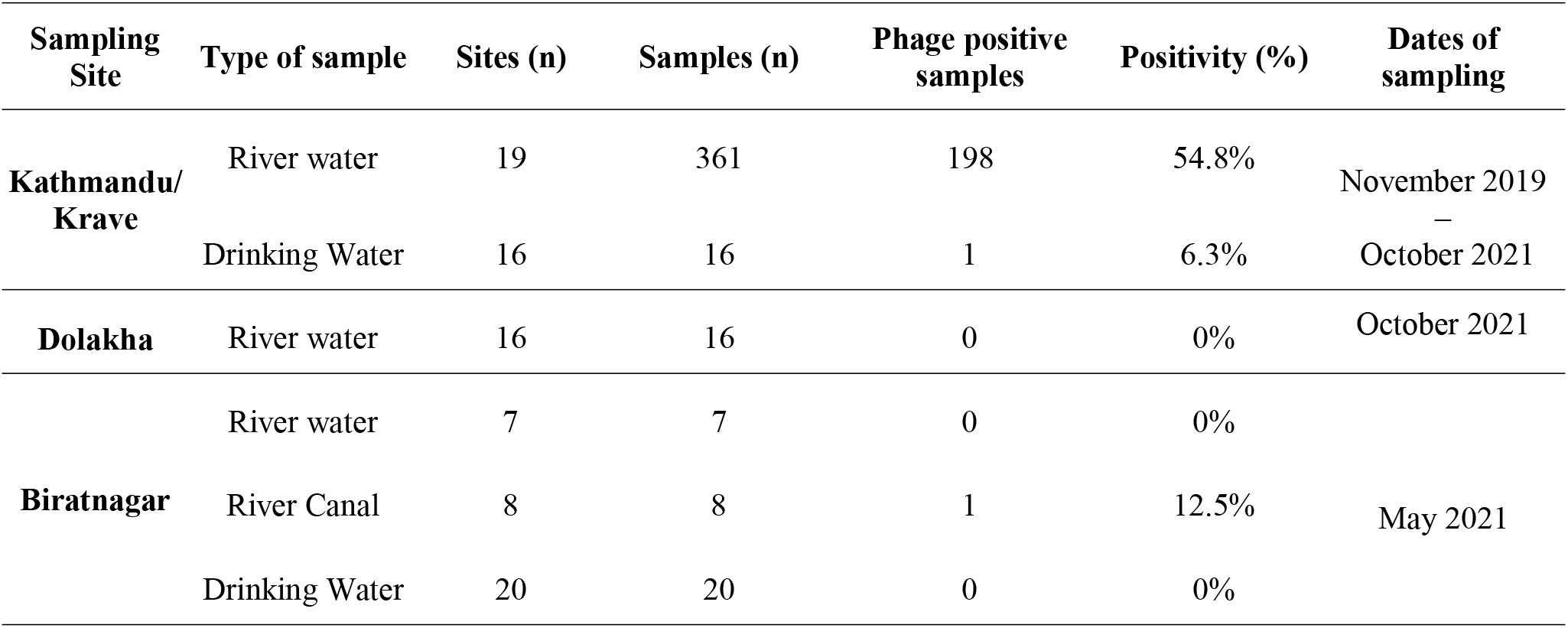
*S*. Typhi phage isolation from environmental samples collected in Nepal.

**Figure 1.**
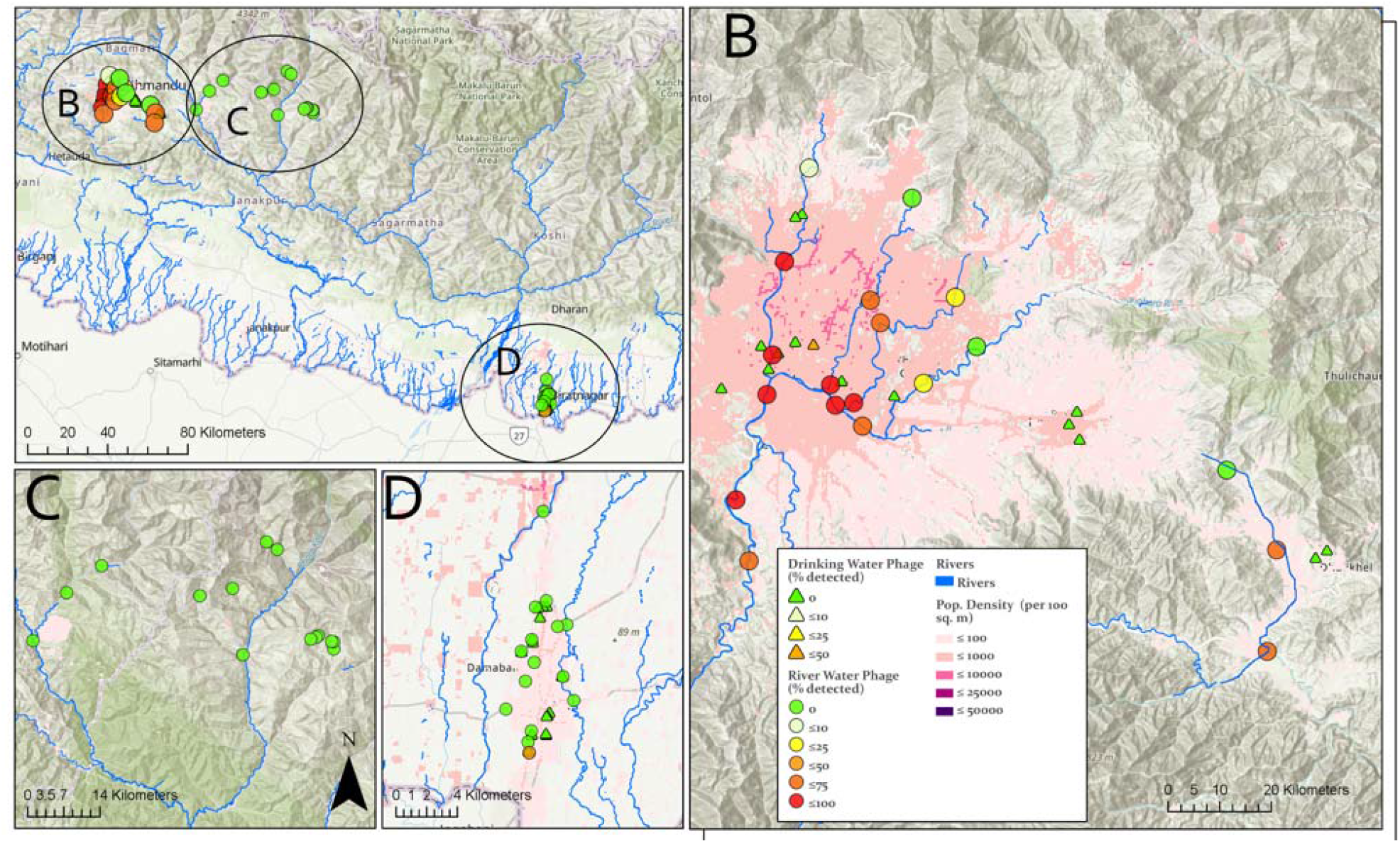
**(a)** Sampling location and detection of *Salmonella* Typhi phages in Nepal. Environmental water sample collection locations and percentage of samples collected with phage detection performed in (**b**) Kathmandu Valley and neighboring Kavrepalanchok district, (c) rural Dolakha and (**d**) urban Biratnagar. Topographic maps are overlayed with population density by shades of red.

### 3.2. Host range properties

To evaluate the host range and specificity of *S*. Typhi phages isolated, we tested a total of 50 phages against a panel of relevant strains including additional *Salmonella enterica* serovars (Paratyphi A and Typhimurium) and other bacteria such as *Pseudomonas aeruginosa, Staphylococcus aureus* and *Klebsiella pneumoniae*. Host ranges were determined by spotting phage solution onto bacterial lawns. The results show that all 50 phages tested were only capable of infecting *S*. Typhi strains and could not infect other bacteria. Although a flagella knockout strain (Δ*fliC*) was susceptible to phage infection, no plaques were formed on the Ty2 isogenic Vi-negative *S*. Typhi strain (Δ*tviB*). Additionally, to understand the diversity of the phages collected, we tested the host range activity of 10 phages against a panel of 37 *S*. Typhi strains belonging to 14 different genotypes circulating in Nepal (Figure 2). The phages tested were isolated from different sampling sites at different time points and were randomly selected, based on different plaque morphologies. As expected, the results showed a broad lytic activity against the *S*. Typhi strains tested. Interestingly, *S*. Typhi strains belonging to lineages 2.1.7, 3.3.1 and 3.3.2 were resistant to the majority of phages tested.

**Figure 2.**
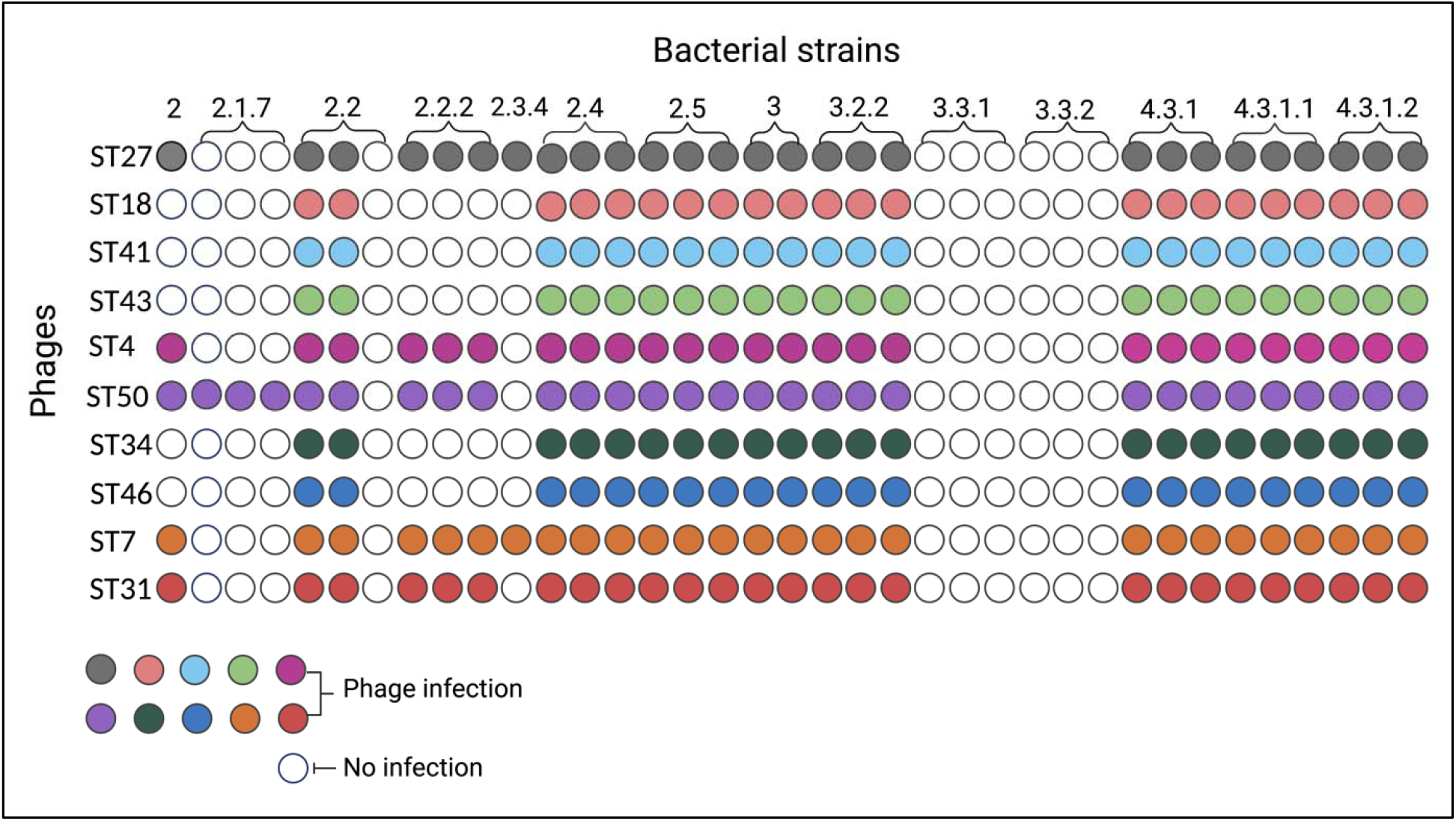
Host range activity of 10 *S*. Typhi phages isolated in our study. *S*. Typhi phages (y-axis) were spotted on *S*. Typhi isolates (x-axis) belonging to different genotypes. Colored circle represent lytic activity, white circles represent no lytic activity.

### 3.3. Characterization of phage genomes

The basic genome metrics of the 27 phage sequences included in this study (host, genome size, GC content, number of predicted ORFs) are provided in Table 1. The analyzed *S*. Typhi phages showed diversity of genome size from 37 to 47 kb, encoding 44 – 73 ORFs. No tRNAs were predicted in the phage genomes.

**Table 2.** Genome characteristics of 27 *S*. Typhi phages sequenced in our study.

|  | Phage | Family | Genome size (bp) | GC % | CDS |
| --- | --- | --- | --- | --- | --- |
| 1 | ST1 | *Autographiviridae* | 38,461 | 50.9 | 49 |
| 2 | ST3 | *Autographiviridae* | 38,461 | 50.9 | 48 |
| 3 | ST10 | *Autographiviridae* | 38,059 | 51.1 | 53 |
| 4 | ST11 | *Autographiviridae* | 38,664 | 51.1 | 52 |
| 5 | ST15 | *Autographiviridae* | 38,351 | 50.9 | 49 |
| 6 | ST16 | *Autographiviridae* | 38,234 | 50.9 | 51 |
| 7 | ST17 | *Autographiviridae* | 38,668 | 50.9 | 52 |
| 8 | ST20 | *Autographiviridae* | 37,644 | 51.0 | 44 |
| 9 | ST21 | *Autographiviridae* | 38,416 | 51.0 | 48 |
| 10 | ST29 | *Autographiviridae* | 39,065 | 51.0 | 49 |
| 11 | ST34 | *Autographiviridae* | 38,880 | 51.0 | 48 |
| 12 | ST35 | *Autographiviridae* | 38,880 | 51.0 | 48 |
| 13 | ST37 | *Autographiviridae* | 38,880 | 51.0 | 48 |
| 14 | ST38 | *Autographiviridae* | 37,743 | 50.9 | 48 |
| 15 | ST43 | *Siphoviridae* | 45,854 | 46.2 | 72 |
| 16 | ST44 | *Siphoviridae* | 45,854 | 46.2 | 73 |
| 17 | ST45 | *Autographiviridae* | 38,351 | 50.9 | 50 |
| 18 | ST46 | *Siphoviridae* | 47,596 | 46.1 | 76 |
| 19 | ST53 | *Autographiviridae* | 39,134 | 50.9 | 52 |
| 20 | ST55 | *Autographiviridae* | 39,134 | 50.9 | 52 |
| 21 | ST56 | *Autographiviridae* | 37,877 | 51.0 | 50 |
| 22 | ST57 | *Autographiviridae* | 37,407 | 51.0 | 47 |
| 23 | ST59 | *Autographiviridae* | 39,134 | 50.9 | 52 |
| 24 | ST63 | *Autographiviridae* | 39,134 | 50.9 | 52 |
| 25 | ST64 | *Autographiviridae* | 39,133 | 50.9 | 52 |
| 26 | ST65 | *Siphoviridae* | 45,855 | 46.2 | 73 |
| 27 | ST66 | *Autographiviridae* | 39,118 | 50.9 | 53 |

The tail fiber and large terminase subunit protein sequences are commonly used markers for understanding phage phylogeny. The phylogenetic relationship was established using the identified terminase and tail fiber nucleotide sequences. All sequences were aligned, and phylogenetic analysis showed that *S*. Typhi phages isolated in our study belong to two different viral families (*Autographiviridae* and *Siphoviridae*) and form three distinct phylogenetic clusters, with two subgroups for the *Autographiviridae* family were found (Figure 3). To better understand the phylogenetic and evolutionary relationship of the phages from our collection, we compared their genomes with previously sequenced phages deposited in GenBank. A total of 27 representative phage genomes belonging to five different families were included and our analysis identified at least 9 distinct clusters. Our findings showed that phages from our collection did not cluster closely with any other phage previously described.

**Figure 3.**
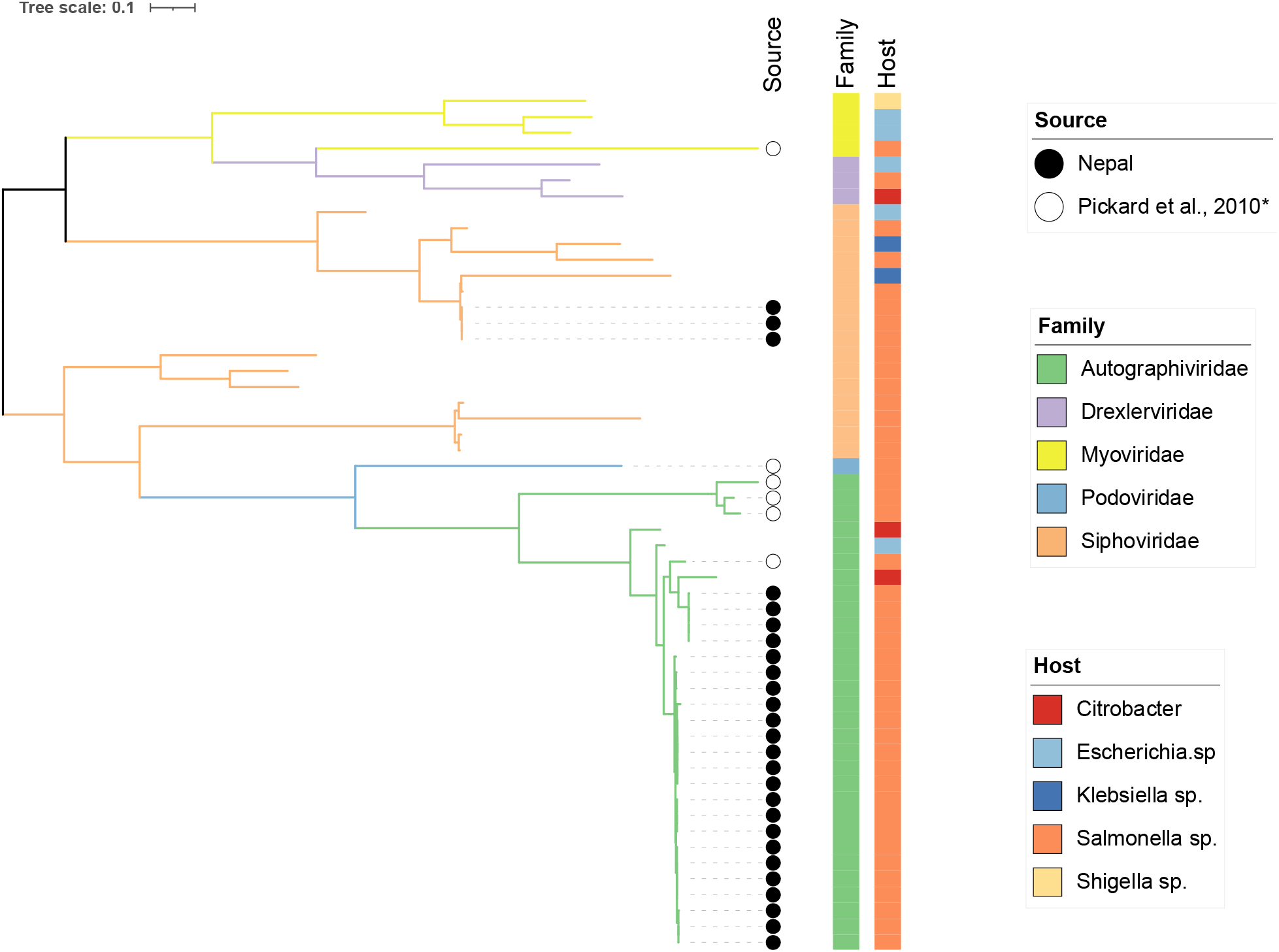
Phylogenetic tree of 27 S. Typhi phages isolated from Nepal and other 27 phages based on two gene sequences (tail fiber and terminase). Tail fiber and terminase large subunit sequences of related phages were downloaded from NCBI. Phages isolated in our study are annotated with a black circle. Pickard et al., 2010* represents a *S*. Typhi Vi phage collection obtained from Cambridge University, Cambridge, United Kingdom, and originally isolated between the 1930 and 1955. These historical phages are annotated with white circles.

## 4. Discussion

In typhoid-endemic communities in and around the Kathmandu Valley of Nepal, we found that *S*. Typhi-specific bacteriophages were widely present in multiple rivers that were contaminated with sewage and readily detectable by small-volume water sampling. By contrast, in a rural and an urban community where typhoid is uncommon, and no visible waste contamination was observed, *S*. Typhi bacteriophages were rarely detected. Taken together, these findings support the premise of phage-based assays as a simple and low-cost tool for environmental surveillance for typhoid, which could be useful for identifying settings to prioritize for vaccination and clean water and sanitation interventions or monitoring their impact after introduction.

Although the detection of *S*. Typhi DNA in municipal drinking water in Kathmandu has been previously reported (22), in our study *S*. Typhi phages were detected in only one drinking water sample. These findings are consistent with our previous work conducted in Kathmandu and Kavrepalanchok, Nepal. We collected drinking water samples and performed quantitative real-time PCR to detected DNA sequences specific for *S*. Typhi. Three hundred and eighty water samples were collected from a randomly selected subset of households and none of these samples tested positive for *S*. Typhi (23).

Our pilot study aimed to generate an easily reproducible approach to be performed in other sites to investigate the role of phages in shaping epidemiologic patterns of typhoid transmission. This will provide an opportunity to elucidate potential differences and similarities in the distribution and abundance of *S*. Typhi in different regions. Our results show that the detection of phages is a cheaper and simpler process than the common methodologies used for detecting bacterial DNA from water samples. Our simplified approach requires minimal supplies, including media for bacterial growth, an incubator, a centrifuge, pipettes, petri dishes, tubes, syringes, and syringe filters. The total cost is <$2 for each sample and given the minimal resources required can be done in labs with very basic infrastructure and equipment.

Our results showed that phages isolated in our study are highly specific to *S*. Typhi isolates. As *Salmonella* consists of more than 2,500 serovars, infection efficacy and specificity are indispensable requirements to the establishment of phage environmental surveillance as a marker for typhoid burden. To further investigate patterns of host infectivity, we tested phages of our collection against Vi capsule-negative and flagellum-negative *S*. Typhi strains. Phages isolated in our study did not infect Vi-capsule mutant strain, demonstrating the requirement of Vi capsule expression for infectivity, as characterized in earlier work (24). We also evaluated the ability of phages to infect different *S*. Typhi genotypes. Interestingly, phages were not able to infect lineages 3.3.1 and 3.3.2., suggesting that some *S*. Typhi lineages can develop phage defense strategies to prevent phage infection and cell lysis. Bacteria can evade phage infection by several mechanisms, including accumulating escape mutations in the receptor, acquiring phage inhibitory proteins, or directly modifying the receptor and using CRISPR-Cas system as a phage defense mechanism (25). Further studies are needed to elucidate the specific host defense mechanisms that *S*. Typhi uses to protect against phage predation, and whether the acquisition of these defense mechanisms drives selection for certain lineages containing them.

Phages from our collection were sequenced and analyzed on the base of genetic similarity to each other and to other phage genomes previously described. Sequence comparison allowed for clear discrimination of distinct clusters. In addition, phylogenetic analysis showed that phages isolated from Nepal clustered independently from other phages included in our analysis; however, a limited number of *S*. Typhi-specific phages have been sequenced and reported. Further studies investigating *S*. Typhi phages in neighboring typhoid-endemic countries and in other regions of the world could further the position of the phages we identified within the global diversity of *S*. Typhi phages.

A series of studies from the cholera field revealed that *Vibrio cholerae* phages exhibit predator-prey dynamics that shape cholera abundance in water, potentially altering seasonal disease patterns (26). A study from Kolkata in 1930 found a similar inverse, seasonal relationship between *S*. Typhi phage abundance and clinical cases, which could support a similar disease dynamic. In our study, seasonal variation was not observed for phage detection. Further studies are needed to understand the potential role of *S*. Typhi phages in shaping the ecology of endemic typhoid transmission.

The results of this study should be interpreted within the context of several limitations. First, we collected a limited number of drinking water samples, and although we observed a low positivity rate, this doesn’t mean that drinking water is not an important transmission route for typhoid in this setting. It is possible that drinking water contamination is more sporadic and wasn’t detected in our limited sampling. Additionally, the scarcity of drinking water in the Kathmandu Valley has led to increased reliance on drinking water that is brought in by large tanker trucks from outside the valley, where typhoid incidence (and contamination risk) is likely lower. Second, the presence of phages in river water reflects their presence in sewage, which is helpful for monitoring burden in the population, but does not directly implicate river water in the transmission pathway. Third, further studies are needed to validate our assay quantitatively in order to specify a limit of detection and sensitivity against a gold standard, such as PCR-based detection methods (9). Fourth, further studies are required correlating frequency, level, geography, and timing of detections of different phages with estimates of disease burden.

Our study highlights the strength of a simplified phage plaque assay as a low-cost tool to identify communities where typhoid is endemic. The high abundance of phages in river water suggest that this could be a scalable and more sensitive alternative to molecular methods for environmental surveillance for typhoid. The findings of our study will inform potential widely implementation of phage environmental surveillance to generate data to support vaccine introduction, monitor intervention strategies and improve understanding of the emergence and transmission of *S*. Typhi in high-risk settings.

## Data Availability

All data produced in the present work are contained in the manuscript

