## Supplementary figure 1 for "Detection of *Salmonella* Typhi bacteriophages in surface waters as a scalable approach to environmental surveillance"

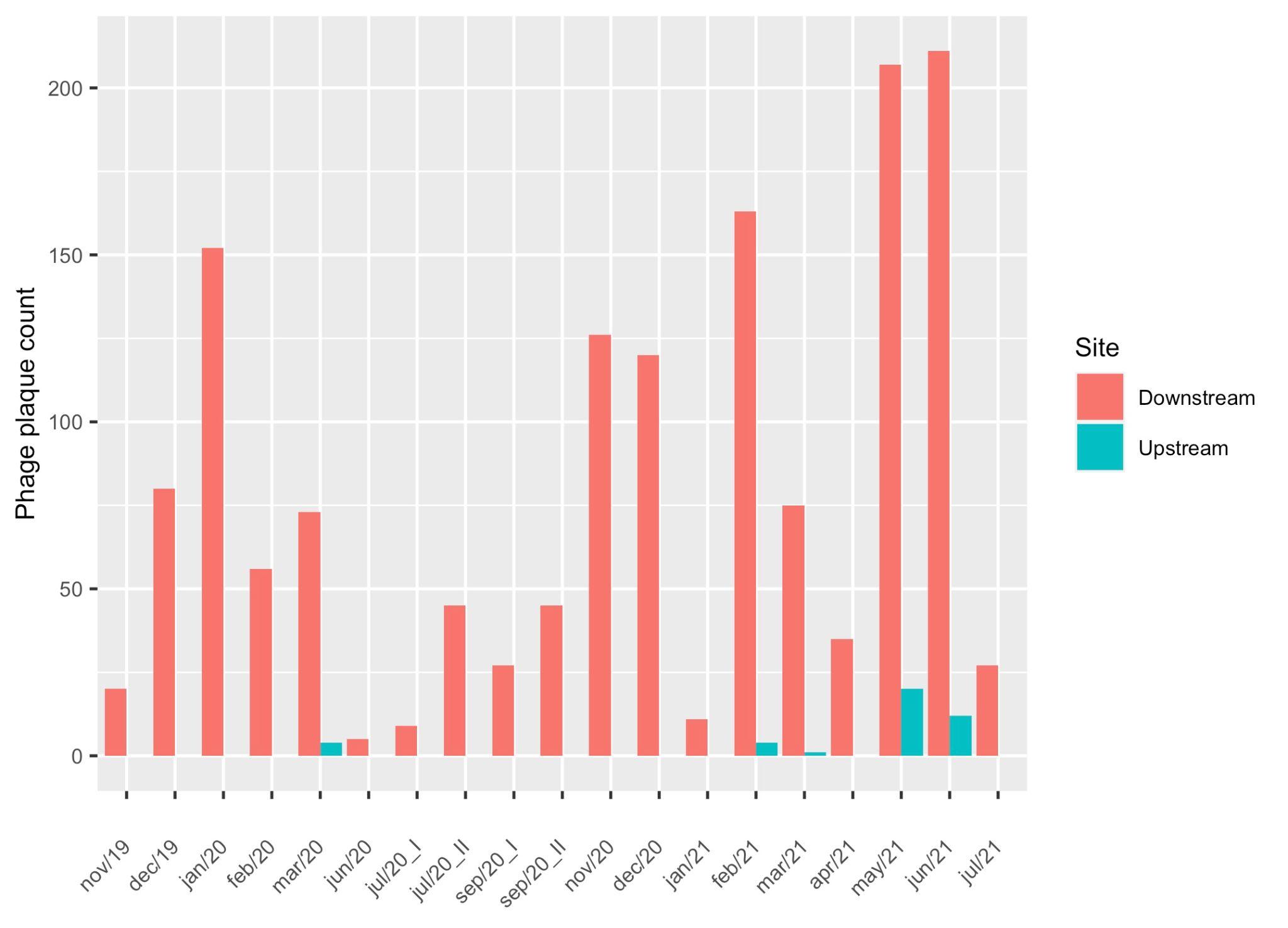

**Supplementary figure 1.** Temporal variation in phage detection in Kathmandu, Nepal during the study period.
